# Trends in accident-related admissions to pediatric intensive care units during the first COVID-19 lockdown in Germany

**DOI:** 10.1101/2021.08.06.21261728

**Authors:** Nora Bruns, Lea Willemsen, Katharina Holtkamp, Oliver Kamp, Marcel Dudda, Bernd Kowall, Andreas Stang, Florian Hey, Judith Blankenburg, Sabir Hemmen, Frank Eifinger, Hans Fuchs, Roland Haase, Clemens Andrée, Michael Heldmann, Jenny Potratz, Daniel Kurz, Anja Schumann, Merle Müller-Knapp, Nadine Mand, Claus Doerfel, Peter Dahlem, Tobias Rothoeft, Manuel Ohlert, Katrin Silkenbäumer, Frank Dohle, Fithri Indraswari, Frank Niemann, Peter Jahn, Michael Merker, Nicole Braun, Francisco Brevis Nunez, Matthias Engler, Konrad Heimann, Gerhard Wolf, Dominik Wulf, Claudia Hollborn, Holger Freymann, Nicolas Allgaier, Felix Knirsch, Martin Dercks, Julia Reinhard, Marc Hoppenz, Ursula Felderhoff-Müser, Christian Dohna-Schwake

## Abstract

**Objective:** To compare the number of accident- and injury-related admissions to pediatric intensive care units (PICU) during the first German COVID-19 lockdown with previous years. To investigate if shifts in types of accidents or injuries occurred, especially regarding non-accidental injuries.

**Design:** Retrospective observational multicenter study.

**Setting:** 37 German PICUs.

**Patients:** 1444 children and adolescents < 18 years admitted to German PICUs due to trauma or injuries during the first German lockdown period (16.3.-31.5.2020) and during the same periods of the years 2017-2019.

**Interventions:** None.

**Measurements and main results:** Standardized morbidity ratios (SMR) and 95% confidence intervals (CI) were calculated for the severity of disease, admission reasons, types of accidents, injury patterns, surgeries and procedures, and outcomes. Disease severity did not differ from previous years. We found an increase in ingestions (SMR 1.41 (CI 0.88 – 2.16)) and a decrease in aspirations (0.77 (0.41 – 1.32)) and burns (0.82 (0.59 – 1.12)). The total number of admissions for trauma remained constant, but traffic accidents (0.76 (0.56 – 1.01) and school/kindergarten accidents (0.25 (0.05 – 0.74) decreased. Household (1.32 (1.05 – 1.64)) and leisure accidents (1.32 (1.05 – 1.65)) increased. Injured structures did not change, but less neurosurgeries (0.69 (0.42 – 1.07)) and more visceral surgeries (2.00 (1.14 – 3.24)) were performed. Non-accidental non-suicidal injuries declined (0.85 (0.50 – 1.37)). Suicide attempts increased in adolescent boys (1.57 (0.58 – 3.42)), while there was a decrease in adolescent girls (0.86 (0.53 – 1.31)).

**Conclusions:** Our study showed shifts in trauma types and associated surgeries during the lockdown period that are generally in line with current literature. The decreased number of non-accidental non-suicidal injuries we observed does not suggest a fundamental increase in severe child abuse during the lockdown period. The decrease in suicide attempts among adolescent girls confirms previous findings, while the increase among boys has not been described yet and deserves further investigation.

## Introduction

By the beginning of March 2020, the spread of the new corona virus SARS-CoV-2 had reached most parts of Europe including Germany. To prevent an uncontrolled transmission of the virus, the state and federal governments announced drastic restrictions to public and private life. The lockdown came into effect on March 16th and restrained many of children’s daily activities: Schools, kindergartens, and sports facilities closed; children’s private contacts had to be reduced to one single friend. This first lockdown to control the pandemic lasted until the end of May 2020, when the restrictions were gradually loosened.

Several authors have reported drastic declines in pediatric emergency department visits, which were mainly driven by declines in infectious diseases (1–3). In the Netherlands and New Zealand, less children presented to emergency departments with trauma (4, 5). At the same time, the lockdown with restrictions to public and private life posed a completely new and unknown challenge for families in Germany. Parents had to work at home and simultaneously care for their children, which lead to psychosocial stress (6, 7). High rates of clinical anxiety and depression among parents have been reported from the United States during this time (8). Across countries, there was major public concern that the lockdown restrictions would lead to an increase in unrecognized child abuse due to lack of social control by schools and kindergartens.

The aim of this study was to compare the number of accident- and injury-related admissions to pediatric intensive care units (PICU) during the German COVID-19 lockdown compared to previous years and to investigate if shifts in types of accidents or injuries occurred compared to previous years. To address this issue, we conducted a retrospective observational multi-center study among 37 Pediatric Intensive Care Units (PICU) across the country. We decided to study only PICU admissions because we reasoned that children with severe injuries would be admitted to the PICU regardless of reduced emergency department visits due to lockdown restrictions.

## Methods

### Study design and recruitment

The study was designed as a retrospective observational multicenter analysis. Members of an informal mailing list of the German Society of Neonatal and Pediatric Intensive Care (GNPI) were inquired via email to participate in the study. Additionally, German children’s hospitals with intensive care units were identified via the homepage of German Society for Pediatrics (Deutsche Gesellschaft für Kinder-und Jugendmedizin). Inquiries for participation in the study were sent out to representatives via email twice between September of 2020 and February of 2021.

### Eligibility and identification of cases

Patients < 18 years of age admitted to a German pediatric intensive care unit due to trauma or injuries during the period of the German COVID-19 lockdown (March 16^th^ – May 31^st^) in the years 2017 – 2020 were eligible. The period of interest was 2020, the years 2017-2019 served as reference period. Eligible diagnoses were S00 – S99 and T00 – T78 according to the German modification of the ICD system (ICD-10-GM) of the corresponding years. S codes apply for trauma diagnoses and T codes apply for other types of injuries or damage from external sources. No code changes were made to the diagnoses of interest between 2017 and 2020. Eligible patients were identified through local hospitals’ medical controlling.

### Data acquisition

Anonymized clinical data were extracted from discharge summaries. Data were entered online into a questionnaire hosted at Microsoft Office Forms 365 for institutional users by the participating centers themselves. Alternatively, anonymized discharge letters were mailed or emailed to the principal study site (Department of Pediatrics I, the University Hospital Essen) and entered by local staff (LW and KH). After the completion of data collection, the raw data were downloaded as a Microsoft Office Excel file and imported into SAS Enterprise Guide 8.4 for statistical analyses.

### Statistical analyses

Continuous variables are presented as median with interquartile range (IQR) and mean with confidence intervals (CI). For discrete variables, absolute and relative frequencies are given. Standardized morbidity ratios (SMR) for the lockdown period were calculated adjusting for age and sex. The years 2017-2019 served as reference period to calculate the expected number of cases for 2020. The observed number of cases in 2020 was then divided by the expected number of cases. An SMR > 1 indicates an increase in cases, an SMR < 1 a decrease. We calculated exact 95% confidence intervals (CI) if the number of observed events in the lockdown period was < 15 and employed the Poisson approximation in case of ≥ 15 events (9).

Age groups for calculation of SMRs were defined as 0-1 years, 2-5 years, 6-11 years, and 12-17 years. Three patients with diverse gender, all from the reference period, were excluded from SMR calculations because no patient with diverse gender was admitted during the observation period.

SAS Enterprise Guide 8.4 (SAS Institute Inc., Cary, NC, USA) was used to perform statistical analyses and produce figures. SISA software (10) was used to calculate exact and Poisson CIs for SMRs.

### Ethics approval

The study was approved by the ethics committee of the Medical Faculty of the University of Duisburg-Essen (20-9560-BO). Local ethics committees of the participating centers additionally approved of the study if required by local legislation. Data entry and storage in Microsoft Office Forms is in line with the General Data Protection Regulation of the European Union (*Regulation (EU) 2016/679*).

## Results

We recruited 37 German PICUs (18 of these University Hospitals) that contributed 1483 cases. 31 cases were excluded because the inclusion criteria were not fulfilled. 8 cases with T diagnoses were excluded from analysis because they were not accidents or injuries. This happened because some subcategories of the ICD codes T75, T76, and T78 include non-injury diagnoses.

The remaining 1444 cases were analyzed. In the reference period, 1098 cases were admitted, averaging 366 cases per year. During the lockdown period, 346 cases were admitted. The median number of reported cases per hospital was 52 (interquartile range 19 - 118). Clinical details of the included patients are given in table 1.

**Table 1:** Patients, admission, and interventions

|  | Overall<br>n (%) | reference period<br>(2017 – 2019)<br>n (%) | 2020<br>n (%) |
| --- | --- | --- | --- |
| Admissions (n) | 1444 | 1098 | 346 |
| Admissions < 1 year of age [n (%)] | 121 (8.4 %) | 91 (8.3 %) | 30 (8.7 %) |
| Age [median (IQR)<br>mean (95% CI)] | 6.5 (2 – 13)<br>7.5 (7.2 – 7.8) | 7 (2 – 14)<br>7.6 (7.3 – 8.0) | 6 (2 – 12)<br>7.3 (6.7 – 7.8) |
| Male [n (%)] | 787 (54.5 %) | 588 (53.6 %) | 199 (57.5 %) |
| Female [n (%)] | 654 (45.3 %) | 507 (46.2 %) | 147 (42.5 %) |
| Diverse [n (%)] | 3 (0.2 %) | 3 (0.3 %) | 0 (0 %) |
| Length of stay on intensive care unit (days)<br>[median (IQR)<br>mean (95% CI)] | 2 (1 – 3)<br>3.4 (3.0 – 3.8) | 2 (1 – 3)<br>3.5 (3.1 – 3.9) | 2 (1 – 3)<br>3.2 (2.3 – 4.0) |
| Mechanical ventilation [n (%)] | 259 (17.9 %) | 198 (18.0 %) | 61 (17.6 %) |
| Duration of mechanical ventilation (days)<br>[median (IQR)<br>mean (95% CI)] | 1 (1 – 3)<br>3.3 (2.5 – 4.1) | 1 (1 – 3)<br>3.7 (2.6 – 4.7) | 1 (1 – 2)<br>2.2 (1.5 – 2.8) |
| Vasopressors [n (%)] | 101 (7.0 %) | 78 (7.1 %) | 23 (6.7 %) |
| Resuscitation [n (%)] | 48 (3.3 %) | 38 (3.5 %) | 10 (2.9 %) |
| Anticonvulsives [n (%)] | 54 (3.7 %) | 44 (4.0 %) | 10 (2.9 %) |
| Died [n (%)] | 13 (0.9 %) | 9 (0.8 %) | 4 (1.2 %) |
| Time to death (days) [median (IQR)<br>mean (95% CI)] | 3 (1 – 3)<br>4.1 (3.0 – 3.8) | 3 (1 – 3)<br>3.6 (1.1 – 6.0) | 2.5 (1.5 – 9)<br>5.3 (0 – 15.7) |
| Poor outcome (GOS 1 or 2) [n (%)] | 15 (1.0 %) | 11 (1.0 %) | 4 (1.2 %) |
| Type of accident |  |  |  |
| Aspiration [n (%)] | 68 (4.7 %) | 55 (5.0 %) | 13 (3.8 %) |
| Burn/scalding [n (%)] | 200 (13.8%) | 159 (14.5 %) | 41 (11.9 %) |
| Drowning/suffocation [n (%)] | 16 (1.1 %) | 11 (1.0 %) | 5 (1.5 %) |
| Ingestion [n (%)] | 69 (4.8 %) | 48 (4.4 %) | 21 (6.1 %) |
| Intoxication [n (%)] | 272 (18.8 %) | 220 (20.0 %) | 52 (15 %) |
| Inhalation of toxic gas [n (%)] | 4 (0.3 %) | 3 (0.3 %) | 1 (0.3 %) |
| Electrical injury [n (%)] | 10 (0.7 %) | 9 (0.8 %) | 1 (0.3 %) |
| Trauma [n (%)] | 802 (55.5 %) | 590 (53.7 %) | 212 (61.3 %) |
| Traffic [n (%)] | 235 (29.3 %) | 187 (31.7 %) | 48 (22.6 %) |
| Household [n (%)] | 260 (32.4 %) | 180 (30.5 %) | 80 (37.7 %) |
| Window lintel [n (%)] |  |  |  |
| Leisure/sports [n (%)] | 258 (32.2 %) | 178 (30.2 %) | 80 (37.7 %) |
| School/kindergarten/work [n (%)] | 38 (4.7 %) | 35 (5.9 %) | 3 (1.4 %) |
| Unknown trauma [n (%)] | 11 (1.4 %) | 10 (1.7 %) | 1 (0.5 %) |
| Unknown accident [n (%)] | 3 (0.2 %) | 3 (0.3 %) | 0 (0.0 %) |
| Non-accidental injury [n (%)] |  |  |  |
| Confirmed [n (%)] | 223 (15.4 %) | 182 (16.6 %) | 41 (11.8 %) |
| Confirmed and suspected [n (%)] | 282 (19.5 %) | 226 (20.6 %) | 56 (16.2 %) |
| Non-accidental non-suicidal | 87 (6.0 %) | 70 (6.4 %) | 17 (4.9 %) |
| Suicide attempt |  |  |  |
| Confirmed [n (%)] | 158 (10.9 %) | 129 (11.8 %) | 29 (8.4 %) |
| Confirmed and suspected [n (%)] | 184 (12.7 %) | 145 (13.2 %) | 39 (11.3 %) |
| Injury pattern |  |  |  |
| Skeletal injury [n (%)] | 305 (21.1 %) | 231 (21.0 %) | 74 (24.3 %) |
| Head injury [n (%)] | 483 (33.4 %) | 363 (33.0 %) | 120 (34.7 %) |
| Visceral organ injury [n (%)] | 193 (13.4 %) | 143 (13.0 %) | 50 (14.5 %) |
| Soft tissue injury [n (%)] | 559 (38.7 %) | 415 (37.8 %) | 144 (41.6 %) |
| Surgeries and procedures |  |  |  |
| Skeletal surgery [n (%)] | 140 (9.7 %) | 106 (9.7 %) | 34 (9.8 %) |
| Head surgery (non-neurosurgery) [n (%)] | 44 (3.1 %) | 36 (3.3 %) | 8 (2.3 %) |
| Neurosurgery [n (%)] | 108 (7.5 %) | 88 (8.0 %) | 20 (5.8 %) |
| Soft tissue procedures/surgery [n (%)] | 265 (18.4) | 203 (18.5 %) | 62 (17.9 %) |
| Visceral organ surgery [n (%)] | 39 (2.7 %) | 23 (2.1 %) | 16 (4.6 %) |
| Thoracic drain placement [n (%)] |  |  |  |
| Endoscopy (after aspiration/ingestion (n = 137)) [n (%)] | 92 (67.2 %) | 66 (64.1 %) | 26 (76.5 %) |
| Any procedure/surgery [n (%)] | 494 (34.2 %) | 373 (34.0 %) | 121 (35.0 %) |

Standardized morbidity ratios revealed shifts in causes of admissions (Figure 1). Drowning accidents (SMR 1.29 (95 % CI 0.42 – 3.02) and ingestions (1.41 (0.88 – 2.16)) increased, whereas aspirations (0.77 (0.41 – 1.32)) and burns (0.82 (0.59 – 1.12)) decreased. Admissions due to intoxications (0.92 (0.70 – 1.22)) and trauma did not change (1.07 (0.93 – 1.22), respectively), but within the type of trauma we observed increases in household (1.32 (1.05 – 1.64)) and leisure accidents (1.32 (1.05 – 1.65)) that were accompanied by decreases in school/kindergarten (0.25 (0.05 – 0.74)) and traffic accidents (0.76 (0.56 – 1.01)).

**Figure 1:**
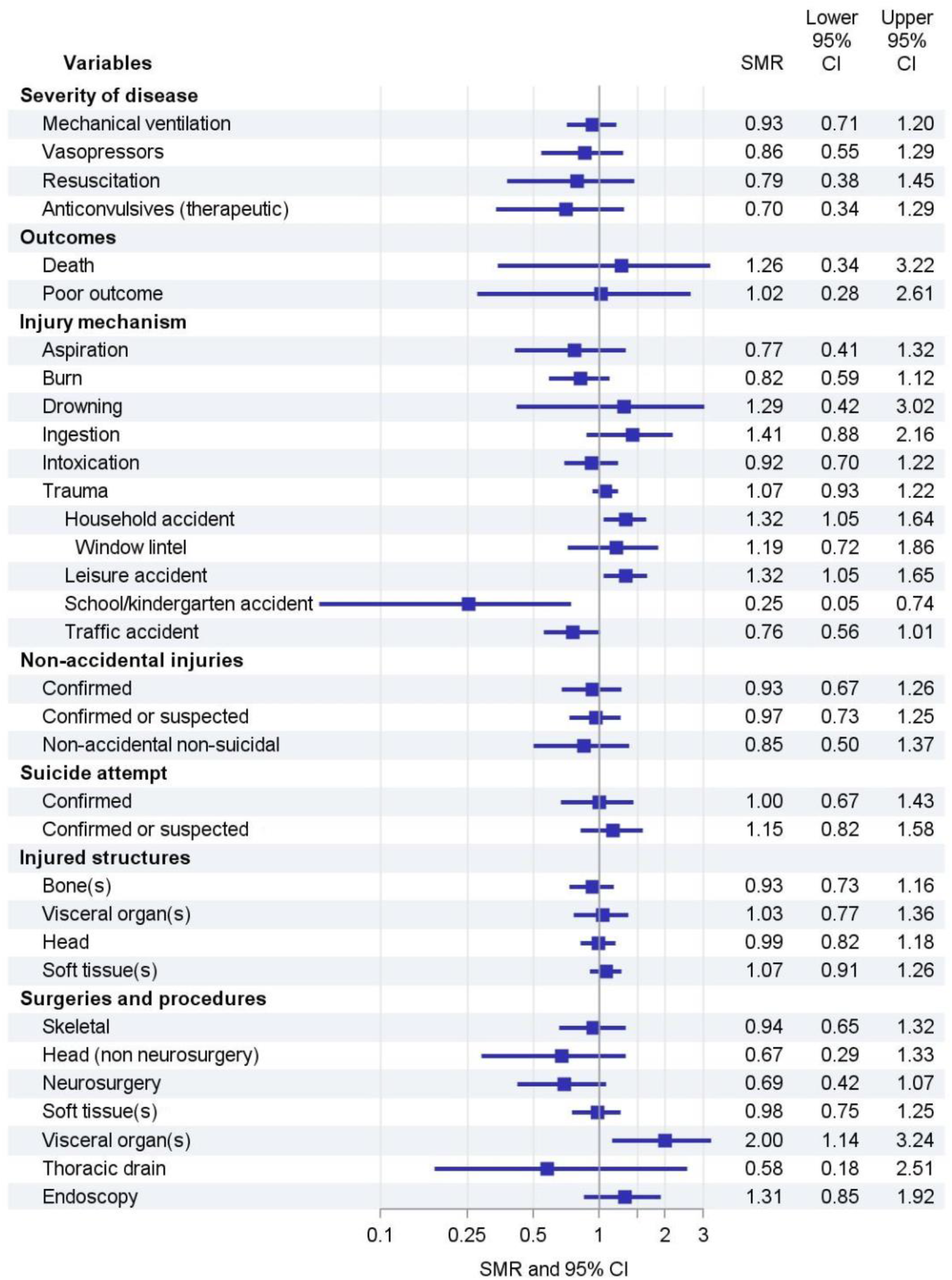
Trends in severity of disease, admissions, injury patterns, procedures, and outcome during the first German COVID-19 lockdown compared to the same periods of 2017-2019. Estimates are presented as standardized morbidity ratios and 95% confidence intervals.

We observed no changes in the injured structures. However, the procedures and surgeries that were performed changed: Surgeries to the head (non-neurosurgery) (0.67 (0.29 − 1.33)) and neurosurgeries (0.69 (0.42 – 1.07)) declined, while at the same time surgeries to visceral organs (2.00 (1.14 – 3.24)) and endoscopies after ingestions or aspirations (1.31 (0.85 − 1.92)) increased.

The SMR for non-accidental injuries did not increase during the study period. The overall SMR for confirmed suicide attempts was 1.00 (0.67 – 1.43), but when confirmed attempts were added up with suspected suicide attempts, the SMR was 1.15 (0.82 – 1.58). Stratification showed that confirmed suicide attempts increased only in adolescent males (SMR 1.57 (0.58 – 3.42)) and decreased in adolescent females (SMR 0.86 (0.53 – 1.31)) (Figure 2). Suspected and confirmed suicide attempts remained stable in adolescent girls but increased in adolescent boys (SMR 1.51 (0.65 – 2.98)) (Figure 2). Suicide attempts at ages < 12 were reported in 2 cases during the reference period and 2 cases during the lockdown and were part of extended suicide attempts.

**Figure 2:**
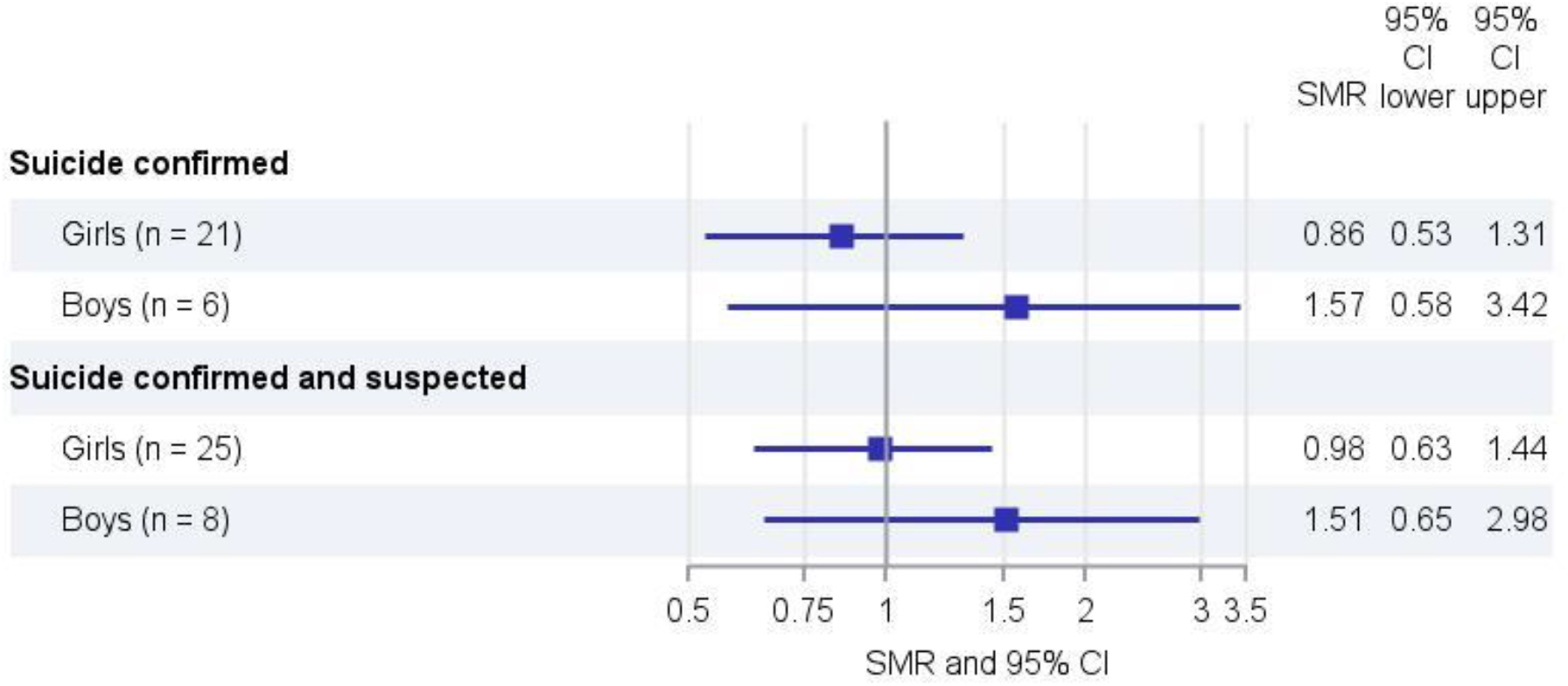
Trends in confirmed and suspected suicide attempts in adolescents during the first German COVID-19 lockdown compared to the same periods of 2017-2019. Estimates are presented as standardized morbidity ratios and 95% confidence intervals.

## Discussion

This multi-center study shows that, despite stable total accident- and injury-related PICU admission numbers during the first German lockdown, there were substantial shifts in the underlying causes of admission. In general, the rates of mechanical ventilation, duration of mechanical ventilation, length of ICU stay, and mortality indicate that patients admitted during the lockdown were not sicker than in previous years. However, there were changes in the type of trauma and consecutive surgeries, even though the overall distribution of injured structures stayed constant. Among non-accidental injuries, in spite of small absolute numbers, the most striking change was an increase in suicide attempts in adolescent boys.

Studies from several countries with varying SARS-CoV2 infection rates and different measures to control the virus spread have reported reduced health care contacts of children either during the pandemic itself or during lockdown periods. In the United States, pediatric hospital admissions declined to 45% at the nadir compared to pre-pandemic years (11). PICU admissions declined by 48 to 70 % (12). In Scotland, emergency PICU admissions for children requiring invasive mechanical ventilation dropped and children admitted to the PICU were not sicker than in previous years, suggesting that they were not admitted to the hospital later than usual (13). The greatest reduction in PICU admissions was seen for respiratory diseases (12, 13); those for injury, poisoning or other external causes were similar to previous years (13). Our data align with these findings, as the total number of admissions slightly decreased during the lockdown and children were not sicker.

The shifts in types of accidents we observed are in line with a report from Israel, where hospitalizations for home trauma increased in children and adults during the lockdown, while less children and adults were admitted after road traffic accidents (14). A similar trend towards reduced motor vehicle accidents during the lockdown was observed in the United States (15). Our data show a decline in admissions after traffic accidents and performed neurosurgeries, which might be related to each other. An international multicenter study recently showed the same trend for non-elective neurosurgeries in adults in Austria, Switzerland and the Czech Republic, that was partially driven by a decline in traumatic brain injuries (16).

Regarding the major concern that non-accidental injuries would increase during the lockdown we observed no clear trend. Contrarily, admissions due to non-accidental non-suicidal injuries declined. In current literature, declines in child protection referrals have mainly been attributed to decreased detection rates (17, 18). In line with this interpretation, a study from the US found an increase of treatments due to physical child abuse trauma (19). In Germany, a representative survey showed that parent-reported adverse childhood events increased during the pandemic, especially the witnessing of domestic violence and verbal emotional abuse. Twenty-nine percent of parents reported an increase in physical child abuse, while 34 % reported a decrease and 37 % no change (20). In another study in Germany, 6.6 % of women reported child corporal punishment (21). During March/April of 2020, suspected cases of child abuse and neglect decreased by 15% in child abuse ambulatories and by 20 % among inpatients in children’s hospitals compared to the same period of 2019 (22). Our data give information only about the most severe cases of non-accidental injuries, as only a small fraction of patients with non-accidental injuries is admitted to pediatric intensive care. Based on the assumption that cases requiring intensive care would have been detected regardless of lockdown restrictions, our data do not support, but neither refute the concern that severe child abuse increased during the lockdown.

Another serious concern that has received international attention is children’s and adolescents’ mental health during the pandemic. Studies showed a high prevalence of depression, anxiety, sleep disorders, post-traumatic stress symptoms, and decreased life satisfaction among children and adolescents during lockdown periods (6, 23, 24). In Italy, the rate of pediatric emergency department visits for psychiatric causes dropped less than the overall rate of ED visits (46% vs. 72%) (25). Admissions for suicidal behaviors in children and adolescents dropped by 50% during the lockdown in France (26). The same trend has been observed in young adults, where incidence rate of admissions due to of self-harm dropped by almost 50%, with stronger effects among females (27). Possibly, the lockdown disrupted some of the dimensions that are associated with suicidal behavior in adolescents, such as school harassment, perceived burdensomeness, and lack of belongingness (26, 28). In our analysis, suicide attempts in adolescent girls declined during the lockdown, in line with findings from literature. However, suicide attempts in boys increased, a phenomenon that has not yet been described. Possibly, protective mechanisms like increased feeling of belongingness and social connectedness did not come into effect in boys.

Our study has several limitations. The most serious limitation derives from the German hospital system: The indication for hospital and intensive care unit admission is at the discretion of the attending physician. This study is based on the assumption that no fundamental changes in admission practice occurred in German PICUs during the study period. Furthermore, the study is not population-based, even though many hospitals and most German University Hospitals participated. Therefore, our estimates rely on case numbers rather than incidence rates. Some of the hospitals in our study used F codes (behavioral disorders caused by psychotropic substances) for intoxications. This prevented several cases from being included, likely causing an underestimation of the number of intoxications during both study periods (reference and observation period). Even though we collected a considerable number of cases, in-depth analyses were limited due to small case numbers in subgroups. As a further consequence, the small case numbers are sensitive to random fluctuations, thus leaving uncertainty regarding the true strength of effect.

## Conclusions

This study is the first to show that the pattern of accident- and injury-related PICU admissions changed during the first COVID-19 in Germany. Even though PICU admissions represent only the tip of the iceberg of all accidents and injuries, our findings are generally in line with present literature on lockdown-associated shifts in admissions to pediatric and adult hospitals. The decline in admissions due to non-accidental non-suicidal injuries is in line with observations from ambulatories and children’s hospitals in Germany. Even though our data do not suggest a dramatic increase in severe child abuse, it cannot be ruled out that the reduced referrals rates to child protection centers were nonetheless a result of decreased detection by school and kindergarten staff. The trend towards lower suicides in adolescent girls aligns with international reports, but the increase among adolescent boys should entail further investigations to verify this observation and, if applicable, offer additional support to this group.

## Data Availability

The dataset generated for this study will be made available to any qualified researcher upon reasonable request.

## Conflicts of interest

The authors have disclosed that they have no conflicts of interest.

## Acknowledgements

None.

## Notes

### Competing Interest Statement

The authors have declared no competing interest.

### Funding Statement

The study received funding from the Stiftung Universitaetsmedizin Essen. NB received funding from the Medical Faculty of the University of Duisburg-Essen (IFORES program) and from the Stiftung Universitaetsmedizin Essen.

### Author Declarations

The study was approved by the ethics committee of the Medical Faculty of the University of Duisburg-Essen (20-9560-BO). Local ethics committees of the participating centers additionally approved of the study if required by local legislation. Data entry and storage in Microsoft Office Forms is in line with the General Data Protection Regulation of the European Union (Regulation (EU) 2016/679).

